# Lack of evidence for interactions between *APOE* and *Klotho* genotypes on cognitive, dementia and brain imaging metrics in UK Biobank

**DOI:** 10.1101/2021.01.11.20248318

**Authors:** Rachana Tank, Joey Ward, Daniel J. Smith, Kristin E. Flegal, Donald M. Lyall

**Author notes:** Corresponding author Dr. Donald M. Lyall, 1 Lilybank Gardens, Institute of Health and Wellbeing, University of Glasgow, G12 8RZ, Glasgow, UK.

## Abstract

**Importance:** Recent research has suggested that genetic variation in the *Klotho* (KL) locus may modify the association between apolipoprotein e (*APOE*) e4 genotype and cognitive impairment.

**Objective:** Large-scale testing for associations and interactions between *KL* and *APOE* genotypes vs. risk of dementia (n=1,570 cases), cognitive abilities (n=174,513) and brain structure (n = 13,158) in older (60+ years) participants.

**Design, setting and participants:** Cross-sectional and prospective data (UK Biobank).

**Main outcomes and measures:** *KL* status was indexed with heterozygosity of the rs9536314 polymorphism (vs. not), in unrelated people with vs. without *APOE* e4 genotype, using regression and interaction tests. We assessed non-demented cognitive scores (processing speed; reasoning; memory; executive function), multiple structural brain imaging, and clinical dementia outcomes. All tests were corrected for age, sex, assessment centre, eight principal components for population stratification, genotypic array, smoking history, deprivation, and self-reported medication history.

**Results:** *APOE* e4 presence (vs. not) was associated with increased risk of dementia, worse cognitive abilities and brain structure differences. *KL* heterozygosity was associated with less frontal lobe grey matter. There were no significant *APOE/KL* interactions for cognitive, dementia or brain imaging measures (all P>0.05).

**Conclusions and relevance:** We found no evidence of *APOE/KL* interactions on cognitive, dementia or brain imaging outcomes. This could be due to some degree of cognitive test imprecision, generally preserved participant health potentially due to relatively young age, type-1 error in prior studies, or indicative of a significant age-dependent KL effect only in the context of marked AD pathology.

**Key points:**

- **Question**: *Klotho* genotype has been previously shown to ‘offset’ a substantial amount of the *APOE* e4/cognitive impairment association. Is this modification effect apparent in large-scale independent data, in terms of non-demented cognitive abilities, brain structure and dementia prevalence?
- **Findings**: In aged 60 years and above participants from UK Biobank, we found significant associations of *APOE* and *Klotho* genotypes on cognitive, structural brain and dementia outcomes, but no significant interactions.
- **Meaning**: This could reflect somewhat healthy participants, prior type 1 error or cognitive/dementia ascertainment imprecision, and/or that *Klotho* genotypic effects are age and neuropathology dependent.

## Introduction

Preserving cognitive abilities such as memory is a common concern into older age, and in the absence of reliable treatments, the public health priority is prevention and delay of cognitive impairment^1^, including understanding effect modifiers. *APOE* e4 is a known risk factor for AD and cognitive decline^1^. Genetic variation in the *KL* locus has been associated with ageing-related phenotypes including insulin resistance and brain function^2^. A recent study of AD cohorts, longitudinal conversion and amyloid-beta samples showed a statistically significant ‘modification’ effect where the deleterious effects conferred by *APOE* e4 were offset by heterozygosity based on two KL polymorphisms in strong linkage disequilibrium: F352V (rs9536314) and C370S (rs9527025), possibly due to correlations with increased serum Klotho^3^. UK Biobank is a relatively large general population cohort^4^ where we have previously shown deleterious effects of *APOE* e4 on cognitive^5^, structural brain imaging^6^ and AD/dementia phenotypes^7^. This brief report tested the hypothesis that based on recent research, genetic KL variation would interact with *APOE* e4 genotype in relevant cognitive, brain and dementia phenotypes.

## Methodology

### Study design and participants

UK Biobank is a prospective cohort study including 502,628 participants who attended one of 22 baseline assessment centres from 2006 to 2010, aged 40-70 years^8^. In 2014, MRI scanning of a sub-group of 100,000 participants began, and this is ongoing. This analysis was conducted under generic approval from the NHS National Research Ethics Service (approval letter dated 17th June 2011, ref 11/NW/0382). Written informed consent was obtained from all participants in the study.

### Dementia outcomes

Dementia/AD outcomes were generated using self-report, hospital admission and death record data, with data utilising International Classification of Diseases version 10 (ICD-10 codes). Individuals were designated as cases (“all-cause dementia” or “Alzheimer disease”) if they had indicated either in self-report or hospital/death records – derived by UK Biobank^7,9^. Those coded as missing were designated as controls (i.e. did not self-report dementia, and diagnoses not present in hospital/death records).

### Imaging data

The release of brain MRI data as of July 2020 was used (i.e. approximately 40k). All imaging data used here was processed and quality checked by UK Biobank^10^. We selected imaging phenotypes *a priori* shown to be associated with worse cognitive ability and decline total white and grey volumes adjusted for skull size (WM/GM respectively); log WM hyperintensity volume (WMH); overall hippocampal volume; general factors of fractional anisotropy (FA) and mean diffusivity (MD), and frontal lobe GM (gFrontal)^11^ based on principal components analysis (PCA). Total WM hyperintensity volumes were calculated based on T1 and T2 fluid-attenuated inversion recovery (FLAIR), derived by UK Biobank.

### Cognitive data

Five tests were completed at baseline (2006-2010), of which we examine three here which have shown sufficient intra-participant reliabilities: Pairs-matching 6-pair (memory), verbal-numeric reasoning and log reaction time (processing speed)^12^. We also examined four cognitive tests administered from 2014 onwards. These were: Trail making test a+b (processing speed/executive function) and Digit symbol substitution (executive function) assessed via online follow-up, plus Matrix pattern completion (nonverbal reasoning) and Tower rearranging (executive function) at MRI^11^.

### Genetic data

UK Biobank genotyping was conducted by Affymetrix using a bespoke BiLEVE Axiom array for ∼50,000 participants and the remaining ∼450,000 on the Affymetrix UK Biobank Axiom array. All genetic data were quality controlled by UK Biobank as described by the protocol paper^4^.). *APOE* e4 ‘risk’ genotype presence (vs. non-e4) was genotyped based on rs7412 and rs429358. KL was indexed using rs9536314 where G/T is considered protective (vs. G/G; T/T) and synonymous with KL-VS diplotype heterozygosity^13^.

### Covariates

Participants self-reported their smoking history: current, past or never, medication use for dyslipidaemia, hormone replacement therapy, blood pressure, oral contraceptive or insulin. We excluded participants for whom these data were missing (<5%). Townsend deprivation indices were derived from postcode of residence.

## Statistical analysis

PLINK v1.90 was used for genetic quality controlling and Stata V.14 was used for statistical analyses. We removed participants who reported neurological conditions as described previously^12^. We statistically controlled for: age, sex, Townsend, ever-smoking, genotypic array, baseline/MRI assessment centre, 8 principal components, array, and medication (concurrent to the phenotype under study; dementia outcomes used baseline values). We focussed on participants aged ≥60 years at baseline^3^. We excluded participants with non-white British ancestry, self-report vs. genetic sex mismatch, putative sex chromosomal aneuploidy, excess heterozygosity, and missingness rate >0.1. We accounted for relatedness between participants by removing one random participant in cases where two individuals were 1st cousins or closer. We have previously reported power calculations indicative of >95% confidence to find ‘true’ effect sizes at Cohen’s D = 0.1 (i.e. small) with regard to *APOE* genotype and outcomes in UK Biobank^6^.

## Results

### Descriptives

After exclusions there were baseline N=174,513 (mean age 64.1, SD=2.85) participants; imaging n=13,158 (72.24 years; SD 3.31). For rs9536314 there were 47,593 heterozygotes (G/T; 27%), and N=46,270 (26.63%) possessed an *APOE* e4 allele. There were n=1,570 dementia cases (0.9%) of which n=634 were AD (0.4%).

### Outcomes

Supplementary Table 1 shows associations between *APOE* e4 genotype and multiple worse outcomes: dementia (odds ratio[OR]=3.27 for e4 vs. not), AD (OR=5.06), log Tower rearranging score (beta = -0.07 SDs for e4 vs. not), Matrix Reasoning (−0.11 SDs), frontal lobe GM (−0.05), hippocampal volume (−0.10), overall GM (−0.06), log TMT total time (0.037) and Digit symbol (−0.075; lowest P-value = 0.038). KL heterozygosity (vs. not) was associated with gFrontal only (−0.041 SDs; P=0.043). There were no statistically significant *APOE/KL* interactions. As sensitivity analyses all models were re-run: unadjusted then covariates added incrementally; excluding people with concurrent neurological conditions^12^; and using the full sample (aged <60). These made no difference to the results.

## Discussion

A recent study by Belloy et al.^3^ reported a protective modifying effect of KL heterozygosity on *APOE* e4 genotype’s conferred risk on cognitive impairment and dementia, in a collation of longitudinal, AD and amyloid-beta cohorts totalling N=24,743. Using unrelated UK Biobank data we tested whether a similar effect could be seen in multiple outcomes: AD/all-cause dementia vs. not, and non-demented cognitive and structural brain MRI phenotypes known to underlie cognitive decline. Participants were ≥60 years as per Belloy et al. We identified individual *APOE* and KL genotype/outcome associations but no interactions, against our hypothesis. There could be an underestimation of true effect due to cognitive test imprecision^12^ or generally preserved participant health^11^. No interaction here vs. Belloy et al. could reflect the use of different phenotypes: the original study investigated AD case vs. control status, conversion to impairment and amyloid-beta while this study investigated AD status and non-demented cognitive/brain structure values. UK Biobank derived dementia status largely from ICD codes whereas Belloy et al. used clinical and/or pathological ascertainment. This could suggest that the age-dependent changes in KL expression, and interaction with *APOE* status, manifest only beyond at least moderate AD-related neuropathology (e.g. amyloid or tau)^14^. The null AD case/control interaction could reflect that UK Biobank is relatively healthy and well-educated. It is possible the UK Biobank participants were not sufficiently old; analysis of longitudinal cognitive and brain imaging data (in independent data) is indicative of more pronounced KL heterozygosity effects particularly into later life^15^.

### Limitations

This study did not explore an exhaustive list of structural imaging phenotypes. There is some degree of healthy volunteer bias in UK Biobank participants, and probably more so in participants who returned for imaging. Some imaging participants would have been unable to complete scanning due to contraindications related to poorer health, e.g. pacemakers/stents^11^.

### Summary

A recent relatively large-scale study including cohort, case/control longitudinal and amyloid-beta data showed a significant interaction whereby KL genotype modified the well-known *APOE* e4 and dementia association. Using independent cognitive, structural brain and dementia data, we did not support these prior findings; this could reflect some degree of bias or imprecision in UK Biobank participants or phenotypes, or that the interaction while ‘true’ is contingent on AD-related neuropathology: future studies should investigate this further in deeply-phenotyped cohorts.

## Data Availability

The UK Biobank resource is available to bona fide researchers for health-related research in the public interest. All researchers who wish to access the research resource must register with UK Biobank (https://www.ukbiobank.ac.uk). Analytic scripts are available upon request.

https://www.ukbiobank.ac.uk/enable-your-research/register

## Funding

RT is funded by a Baillie Gifford doctoral student fellowship.UK Biobank was established by the Wellcome Trust medical charity, Medical Research Council, Department of Health, Scottish Government and the Northwest Regional Development Agency. It has also had funding from the Welsh Assembly Government and the British Heart Foundation. DJS is supported by an MRC Pathfinder grant.

## Role of the funder/sponsor

The funders had no role in study design, data collection or management, analyses or interpretation of the data, nor preparation, review or approval of the manuscript.

## Conflict of interest disclosures

None.

## Author contributions

Concept and design: DML

Acquisition, analysis, or interpretation of data: DML, RT.

Drafting of the manuscript: DML, RT.

Critical revision of the manuscript for important intellectual content: All co-authors.

Statistical analysis: DML.

Obtained principal study funding: DML, KF.

## Acknowledgements

This research has been conducted using the UK Biobank resource; we are grateful to UK Biobank participants. Thanks to Dr. Breda Cullen for devising exclusion criteria and Dr. Michael A. Belloy for helpful comments on an earlier draft. This project was completed using UK Biobank application 17689 (PI: DML)..

## Notes

### Competing Interest Statement

The authors have declared no competing interest.

### Author Declarations

Participants provided full informed consent to participate in UK Biobank. This study was covered by the generic ethical approval for UK Biobank studies from the NHS National Research Ethics Service (approval letter dated 17th June 2011, Ref 11/NW/0382). Approval was received from UK Biobank to use the data in the present work (UK Biobank application #17689; PI Lyall).

